# Characterizing mobility patterns and malaria risk factors in semi-nomadic populations of Northern Kenya

**DOI:** 10.1101/2023.12.06.23299617

**Authors:** Hannah R. Meredith, Amy Wesolowski, Dennis Okoth, Linda Maraga, George Ambani, Tabitha Chepkwony, Lucy Abel, Joseph Kipkoech, Gilchrist Lokoel, Daniel Esimit, Samuel Lokemer, James Maragia, Wendy Prudhomme O’Meara, Andrew A. Obala

## Abstract

While many studies have characterized mobility patterns and disease dynamics of individuals from settled populations, few have focused on more mobile populations. Highly mobile groups are often at higher disease risk due to their regular movement that may increase the variability of their environments, reduce their access to health care, and limit the number of intervention strategies suitable for their lifestyles. Quantifying the movements and their associated disease risks will be key to developing intervention strategies more suitable for mobile populations. Here, we worked with four semi-nomadic communities in Central Turkana, Kenya to 1) characterize mobility patterns of travelers from semi-nomadic communities and 2) test the hypothesis that semi-nomadic individuals are at greater risk of exposure to malaria during seasonal migrations than when staying at their semi-permanent settlements. From March-October, 2021, we conducted a study in semi-nomadic households (n=250) where some members traveled with their herd while others remained at the semi-permanent settlement. Participants provided medical and travel histories, demographics, and a dried blood spot for malaria testing before and after the travel period. Further, a subset of travelers was given GPS loggers to document their routes. Four travel patterns emerged from the logger data, Long Term, Transient, Day trip, and Static, with only Long Term and Transient trips being associated with malaria cases detected in individuals who carried GPS devices. After completing their trips, travelers had a higher prevalence of malaria than those who remained at the household (9.2% vs 4.4%), regardless of gender, age group, and catchment area. These findings highlight the need to develop intervention strategies amenable to mobile lifestyles that can ultimately help prevent the transmission of malaria.

## Introduction

Quantifying the relationship between human mobility and disease transmission is critical for developing more effective interventions(1–5). The majority of these studies have focused on either individuals that travel to/from a permanent residential location or mobility and disease transmission patterns that have been generalized to larger geographic areas(1–9). Fewer studies have focused on more mobile individuals, such as those from semi-nomadic populations, who are more difficult to reach and do not follow the general mobility patterns of the larger population. Relative to settled individuals, they are often at higher disease risk due to their regular movement, reduced access to health care, and lack of interventions suitable for their lifestyles(10–14). In some settings, individuals who move regularly to maintain their livelihoods are often exposed to infectious diseases or high transmission environments more frequently than their settled counterparts(15). For example, nomadic communities have been exposed to increased risks of malaria infection when seeking watering holes for their animals because this brings them in contact with mosquito breeding sites in arid areas(11,16). In other settings, mobile populations’ isolation and frequent movement may result in irregular exposure to diseases in circulation within the settled community. For example, the Tuareg nomads have avoided exposure to measles in the past by avoiding markets and wells for months that they associated with the epidemic (17). While their movement reduced their initial exposure, it could render them more susceptible to outbreaks and more severe symptoms in the future due to reduced acquired immunity compounded by low vaccination coverage(11,17,18). Further, eradication of infectious diseases may be challenging if transmission is concentrated in these hard-to-reach and under-served populations. For instance, both smallpox and polio have been reintroduced into settled communities by nomadic populations who were unvaccinated(14,19). Therefore, a better understanding of mobile populations travel patterns and their relationship with disease dynamics would help determine when, where, and who to focus on in intervention strategies and elimination campaigns.

Typically, human movements have been estimated by census data, traffic and travel surveys, flight statistics, night-time satellite images, call data records (CDRs), social media, and personal global positioning systems (GPS)(6,20–24). These methods have been used to study populations that are easy to locate, own cell-phones, and use established travel networks. However, the resulting datasets may not be relevant for characterizing the movements of mobile, remote populations, such as nomadic communities, that are either difficult to reach or intentionally excluded(15,25). Additionally, regular movements motivated by pastoralism and hunting and gathering are not typically captured by general surveys, like censuses taken every 5-10 years, and would likely be aggregated into larger flows of movement between administrative units (i.e., towns, districts, regions) during mobile phone or social media data pre-processing. Thus, specific studies are needed to characterize mobility patterns of uniquely mobile populations. For example, GPS loggers have been used to characterize travel patterns of mobile populations in Lao PDR, Mongolia, and Senegal(26–28). While a large proportion of the world’s mobile populations reside in Africa and some studies have documented the health challenges and general travel patterns of different mobile populations across Africa(10,12,13,16), few studies have quantified the movement patterns and possible relationship with infectious disease transmission.

Turkana is a semi-arid county in north-western Kenya with a sparse population that is 60% semi-nomadic or nomadic(29) (**Fig 1A**). The mobile lifestyles of the Turkana have been studied from anthropological and ecological perspectives(30–32); however, their travel patterns have not been well quantified, largely relying on travelers recounting trip details in surveys or indicating their routes on maps(29). Similarly, the disease dynamics of the mobile Turkana have not been well studied, relying on a few studies that use self-reporting of health complaints and symptoms that could be associated with certain diseases(33,34). While these studies attributed the Turkana’s health complaints (or lack thereof) to their mobile lifestyles, there has yet to be a study directly relating their travel patterns and risk of disease. To define this relationship, we conducted a study of semi-nomadic households across Central Turkana to better understand their mobility patterns and determine if traveling affects their risk for disease exposure. Specifically, we focused on the risk of malaria exposure because it was recently confirmed to be endemic in Central Turkana(35). However, as the majority of the households enrolled for this previous study were settled, malaria exposure in mobile households from this area remains to be characterized. Since nomadic individuals traveling with their herds tend to congregate at water sources, potential mosquito breeding sites, we hypothesized that the Turkana who travel with their herds are more likely to be exposed to malaria than those who remained in the settled communities. If this is true, they may be importing malaria back to their villages of origin. Ultimately, understanding the extent to which mobile populations impact malaria transmission is key for informing elimination efforts by providing insight on how to better tailor surveillance and intervention strategies for these unique populations.

**Fig 1.**
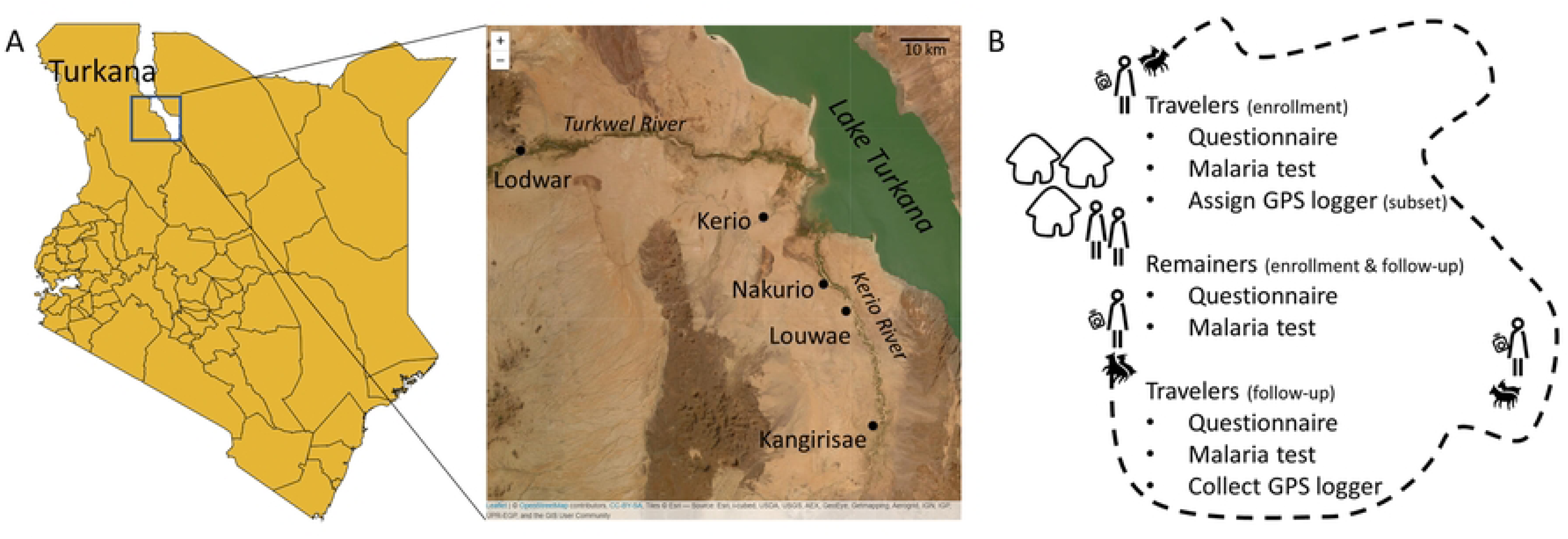
Overview of study area and design. (A) Enrollment took place in Central Turkana (box on left map), near four health facilities (labeled on the right map). (B) Semi-nomadic households with at least one traveler and remainer were enrolled. Before and after travelers migrated with their herds, they provided blood samples for malaria tests and answered questions on recent travel and medical history. GPS loggers were assigned to a subset of those traveling with the herd. Shapefiles were downloaded from DIVA-GIS (https://www.diva-gis.org/) and Esri World Imagery was accessed via the R package leaflet.

## Methods

### Ethic

Written informed consent was provided by all adults and by parents or guardians for individuals under 18 years old. Individuals 12-18 years old were asked for verbal assent. This study was approved by the ethical review boards of Moi University (IREC/2020/209) and Duke University (Pro00107835).

### Study Area

Turkana is a sparsely-populated, semi-arid county in north-western Kenya (**Fig 1A**). Approximately 60% of its residents are (semi-)nomadic(36), relying on their livestock for their livelihood. Despite the sparse population density and very limited rainfall, malaria is endemic in this area (35,37). We enrolled households from semi-permanent settlements in four catchment areas defined by four health facilities in Central Turkana (Kerio, Nakurio, Louwae, and Kangirisae). These facilities are located near a seasonal river (Kerio River) which empties into Lake Turkana to the east. Communities in this area have a large number of households with members who travel seasonally to access food or water for livestock (personal communication with community health workers). While Lake Turkana is nearby, its water is alkaline and typically not used for drinking water (personal communication with community health workers).

### Study Population

We recruited and consented households with at least one person planning to travel with the herd for at least two consecutive weeks (traveler) and at least one person planning to remain behind at the settlement (remainer). Individuals had to be at least one year old to be eligible for participation. At enrollment, before the travelers left with their herds, and again at follow-up, after the travelers returned with their herds, participants provided a finger-prick blood sample for a dried blood spot (DBS) and answered a questionnaire detailing their travel and medical history (**Fig 1B**).

### GPS logger substudy

One traveler per household, either the head of household or the lead herder for the household, was asked to carry a GPS logger during their trip. The number of travelers assigned a GPS logger was limited by both the number of GPS loggers available (48) and when travelers returned to allow for a logger to be reassigned to a new household. The GPS logger (model i-gotU GT-600) was light-weight (< 80 grams), small (46×41.5×14mm), water resistant, battery powered (750 mAh), password protected, has 64 Mb memory with the capability of storing 262,000 location points, and could be worn in multiple ways (i.e., lanyard, velcro, watchband). GPS loggers were programmed to record location, date, and time eight times a day. To conserve battery power and ensure travel patterns were recorded at different times of the day, loggers were programmed to record locations hourly during two moving four-hour windows separated by 12 hours (i.e., Day 1: 12, 1, 2, 3 am and pm; Day 2: 4, 5, 6, 7 am and pm, etc.).

### GPS logger analysis

GPS tracks that covered at least 50% of travel dates, as defined by the dates between enrollment and follow-up, were included in analysis. While the dates of enrollment and follow-up did not always correspond with the departure and return dates reported by travelers, a sensitivity analysis suggests this should not affect the results **(S1 Text)**. To distinguish between short movements around a given point that could be associated with stationary grazing and longer directional movements, sequential GPS points that fell within a 500m radius were hierarchically clustered using the hclust and cutree functions from the geosphere package in R (version 4.2.2). New coordinates based on the centroid and an ID were assigned to each new cluster. Each GPS track was analyzed by plotting the cluster IDs as a function of date and time of day (i.e., night (6pm – 6am) vs day). Night locations were categorized as long-term campsites if a week or more of consecutive nights were spent there or transient campsites if fewer than a week of consecutive nights were spent there (see **S2 Text** for sensitivity analysis).

Travel trajectories from GPS loggers were analyzed individually and then categorized into four ‘trip types’. Long Term trips had the majority of nights logged at long-term camps while Transient trips had the majority of nights logged at short-term camps. Day trips had >90% of night GPS points logged at the same night location recorded on the day they were enrolled, but different GPS points logged during the day. This likely reflects scenarios in which the travelers conducted day trips with their herds and returned to the same camp each night. Static trips had >90% of all points (day and night) logged at the same location, likely representative of scenarios in which the travelers stayed within a 500-meter radius for the duration of the study or the loggers were not carried. GPS points were mapped in R on Esri.WorldImagery provider tiles using the leaflet (version 2.1.1) and leaflet.esri (1.0.0) packages.

### Molecular detection of Plasmodium falciparum

Genomic DNA was extracted from each DBS using the Chelex method. All DBS extracts were screened with genus-specific primers for Plasmodium spp. Positive samples were tested for the presence of *P. falciparum* using species specific primers. The expected product size was 206 bp which was visualized on a 2% agarose gel stained with Sybr safe(38).

### Data capture and statistical analyses

Two community health workers per catchment area (Kerio, Nakurio, Louwae, and Kangirisae) identified and enrolled eligible households, administered the enrollment and follow-up surveys, and collected DBS. Completed paper data collection tools were reviewed monthly with the CHWs to identify and address any data quality issues. Data collected from the surveys were entered into a REDCap database (https://www.project-redcap.org/), and analyzed in R. PCR results from the DBS were saved in Microsoft Excel and imported into R for analysis.

We compared demographic, travel and medical history between the following subpopulations – 1) travelers who did or did not carry loggers to assess generalizability of the logger data, 2) those who traveled or who remained at the settlement and 3) those who did or did not acquire a malaria infection over the travel period. Binomial general regression models were used to quantify correlation between infection outcome and putative risk factors (glm function, R).

## Results

### Traveler Population and Trip Details

Between March and October, 2021, we enrolled 250 households that included at least one person who expected to remain and one who expected to travel (n=929 participants). In total there were 304 members who reported plans to travel with the herd. At follow-up, 70 of these members reported that they had not traveled, 44 additional members reported they had traveled with the herd, and 18 members were lost to follow-up, thus 260 travelers from 249 households were included in the travel analysis (**Fig 2**). The majority of travelers were male (87.7%, 228/260) with a median age of 31 years (interquartile range (IQR) 19-40) and a median trip duration of 57.2 days (IQR 42.2-76.2) (**Table 1**). Of the 260 travelers, 64 carried a GPS logger throughout the entire study period; however, only 58 tracks were analyzed because four GPS did not return data. Two GPS loggers were lost in the field, one lost battery power before the trip started, one did not consistently collect data throughout the trip, and two GPS loggers lasted less than 50% of the travel period.

**Fig 2.**
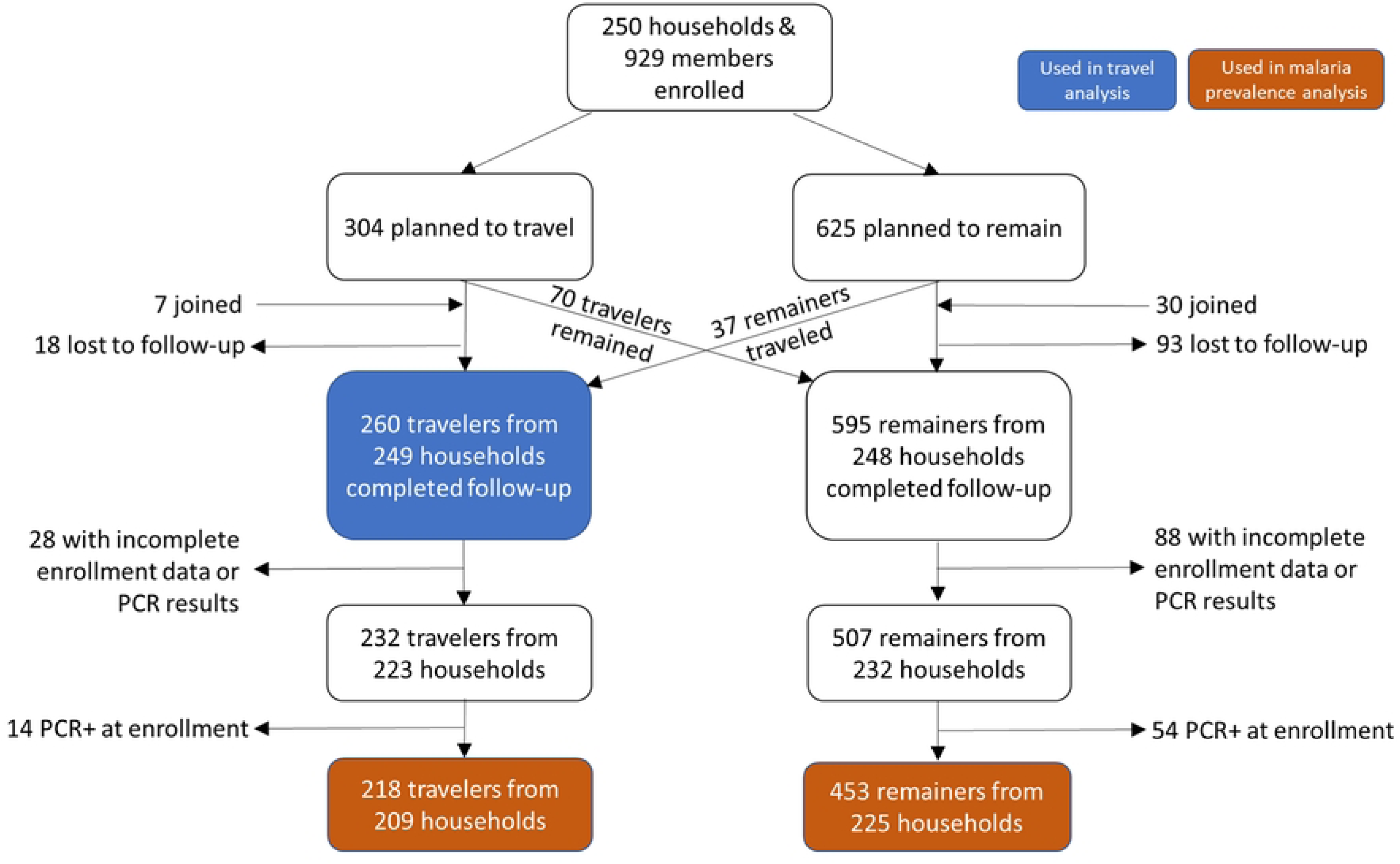
Diagram of inclusion and exclusion for different analyses. For the travel analysis, any traveler who provided trip information was included. For the malaria analysis, remainers and travelers had to provide complete information for both enrollment and follow-up as well as test negative for PCR at enrollment. Accounting for the fact that some households had travelers and/or remainers included in the analysis, there was a total of 242 households represented in the malaria prevalence analysis.

**Table 1.** Characteristics of travelers and trip-types, overall and stratified by GPS carrier status. % (number/N); median (IQR)

|  | All travelers<br>(N =260) | Non-GPS carriers<br>(N =202) | All GPS carriers<br>(N = 58) | GPS trip types |  |  |  |
| --- | --- | --- | --- | --- | --- | --- | --- |
|  |  |  |  | Long term:<br>(N = 24) | Transient:<br>(N = 20) | Daytrips:<br>(N = 3) | Static:<br>(N = 11) |
| **Traveler details** |  |  |  |  |  |  |  |
| Male | 87.7 (228/260) | 86.1 (174/202) | 93.1 (54/58) | 87.5 (21/24) | 100 (20/20) | 100 (3/3) | 90.9 (10/11) |
| Age (years) | 31 (19-40) | 30 (19-40) | 37 (30-45) | 37 (30.0 - 48.5) | 36 (28.8 - 36.0) | 26 (22-44) | 38 (33-46) |
| Catchment area |  |  |  |  |  |  |  |
| Kangirisae | 24.2 (63/260) | 23.8 (48/202) | 25.9 (15/58) | 29.2 (7/24) | 30 (6/20) | 0 (0/3) | 18.2 (2/11) |
| Lowae | 26.2 (68/260) | 26.2 (53/202) | 25.9 (15/58) | 37.5 (9/24) | 5 (1/20) | 33.3 (1/3) | 36.4 (4/11) |
| Nakurio | 25.4 (66/260) | 25.2 (53/202) | 22.4 (13/58) | 8.3 (2/24) | 35 (7/20) | 0 (0/0) | 36.4 (4/11) |
| Kerio | 24.2 (63/260) | 23.8 (48/202) | 25.9 (15/58) | 25 (6/24) | 30 (6/20) | 66.7 (2/3) | 9.1 (1/11) |
| **Reported trip details** |  |  |  |  |  |  |  |
| Trip duration (days) | 57.2 (42.2-76.2) | 57.2 (42.2-75.2) | 57 (39.3-90.3) | 56 (41.5-77.5) | 55 (33.8 - 83.8) | 53 (41-92.5) | 70.0 (47.0-104.5) |
| Camps reported | 1 (1-2) | 1 (1-1) | 1 (1-2) | 1 (1-1.3) | 2 (1-2) | 3 (2-3) | 1 (1-1) |
| Travel time to camp (days) | 2-3 (1:2-3) | 2-3 (1:2-3) | 2-3 (1:2-3) | 2-3 (1:2-3) | 2-3 (1:2-3) | 1-2 (<1:2-3) | 2-3 (2-3:2-3) |
| Non-HH members present | 83.5 (217/260) | 81.7 (165/202) | 89.7 (52/58) | 91.7 (22/24) | 85.0 (17/20) | 66.7 (2/3) | 100 (11/11) |
| People at camp (#) | 4-6 (1-3: 7-10) | 4-6 (1-3: 7-10) | 4-6 (4-6: 7-10) | 4-6 (4-6: 7-10) | 4-6 (4-6: 7-10) | 4-6 (1-3: 7-10) | 4-6 (1-3: 4-6) |
| Nearby water source* |  |  |  |  |  |  |  |
| Open<sup>1</sup> | 86.9 (226/260) | 87.6 (177/202) | 84.5 (49/58) | 83.3 (20/24) | 85.0 (17/20) | 100 (3/3) | 90.9 (10/11) |
| Closed<sup>2</sup> | 30.4 (79/260) | 29.7 (60/202) | 32.8 (19/58) | 42.7 (10/24) | 20 (4/20) | 33.3 (1/3) | 36.4 (4/11) |
| Animals traveled with |  |  |  |  |  |  |  |
| Goats | 99.2 (258/260) | 99.5 (201/202) | 98.3 (57/58) | 100 (24/24) | 95 (19/20) | 100 (3/3) | 100 (11/11) |
| Sheep | 88.0 (229/260) | 88.1(178/202) | 87.9 (51/58) | 87.5 (21/24) | 80 (16/20) | 100 (3/3) | 100 (11/11) |
| Camels | 9.2 (24/260) | 9.4 (19/202) | 8.6 (5/58) | 12.5 (3/24) | 5 (1/20) | 0 (0/3) | 9.1 (1/11) |
| **GPS details** |  |  |  |  |  |  |  |
| Campsite changes | -- | -- | -- | 4 (2 - 8.3) | 17 (11-32.8) | 0 (0-3.5) | 0 (0-1) |
| Campsites logged | -- | -- | -- | 3 (3-5.3) | 10.5 (7.3-18) | 1 (1-3) | 1 (1-2) |
| Total distance between camps (km) | -- | -- | -- | 29.0 (10.6 - 53.2) | 87.8 (69.5 - 210.3) | 1.6 (1.2-34.1) | 2.1 (1.5-5.8) |
| Total distance traveled (km) | -- | -- | -- | 106.8 (35.9 - 156.5) | 278.5 (186.3-557.4) | 157.4 (130.5-186.1) | 33.5 (23.0-54.9) |

\* Water source types were not mutually exclusive – both could be reported by participant. <sup>1</sup>Open water source: River, lake, dam, spring, hand dug water pit; <sup>2</sup>Closed water source: Tap water, well or borehole;

We compared baseline demographic characteristics between those who carried a logger and those who did not to ensure the logger data was representative of travelers more broadly (**Table 1**). The main differences between those who carried loggers and those who did not was the carriers tended to be slightly older (37 years (IQR 30-45) vs 30 years (IQR 19-40)), and had a higher proportion of males (93.1% (54/58) vs 86.1% (174/202)) (**Table 1**). Trip information was similar across groups, although individuals with GPS loggers included a slightly higher proportion of people who reported staying at a campsite with non-household members (89.7% (57/58) vs 81.7% (165/202)) (**Table 1**). Given these similarities, we assumed the GPS loggers carried by a subset of travelers could be generalized as representing the spatial-temporal patterns of this semi-nomadic community’s trips.

### Travel pattern analysis

At the population level, GPS data revealed that travelers from the same catchment area tended to travel in common regions; however, when evaluating the number of different travelers who visited a given point to determine common destinations, most of the points (79.2% of night points and 71.6% of all points) were frequented by a single traveler (**Fig 3A&B, S1 Fig**). “Hot spots”, where multiple travelers were recorded, tended to be near the center of the village or along main corridors of travel (i.e., the route along the Kerio River) (**Fig 3B, S1 Fig**). Overlaying the tracks with satellite imagery showed that, while some points were logged along the Kerio River or the shore of Lake Turkana, many of the trajectories moved away from these larger sources of water.

**Fig 3.**
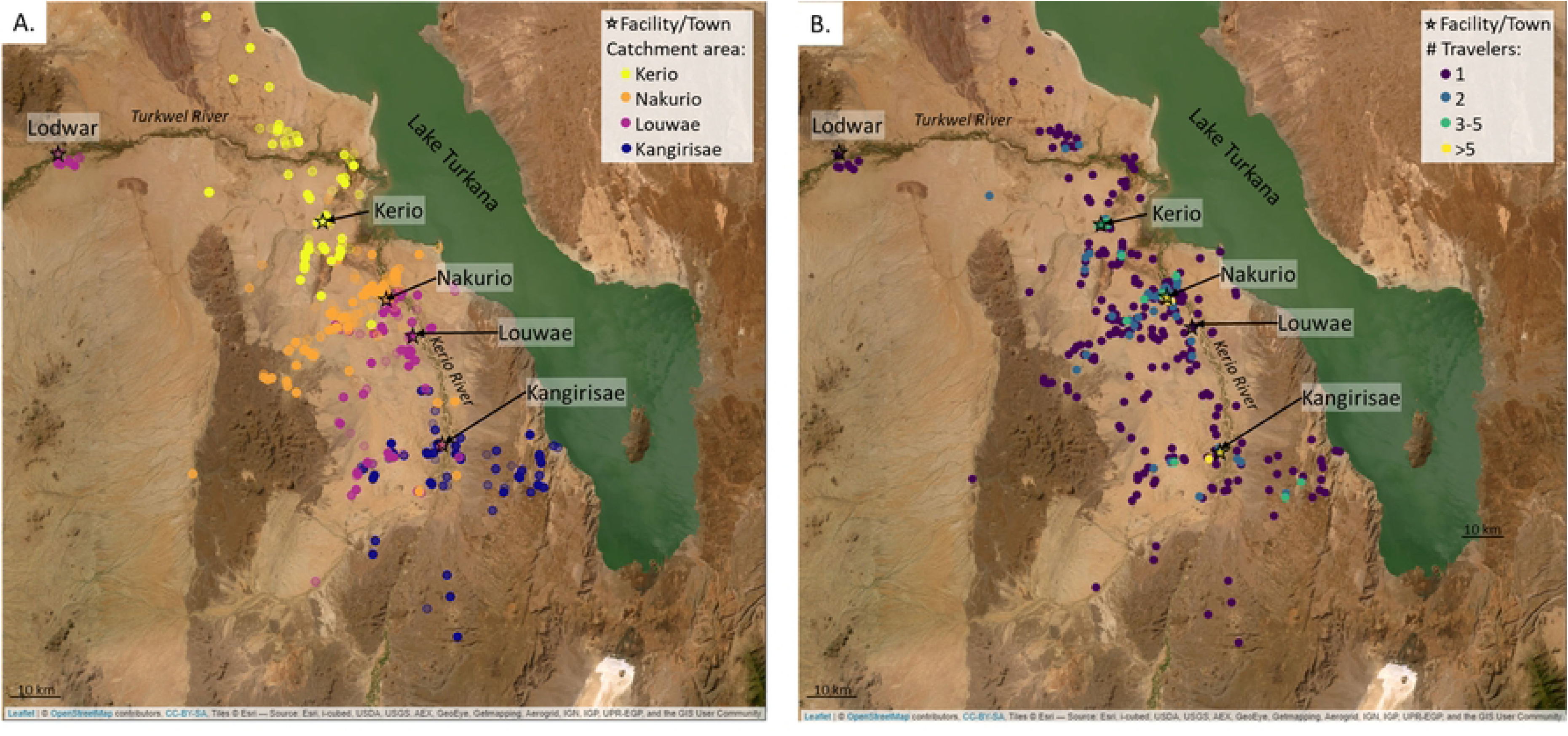
Population level trip characteristics. (A) Campsite locations, stratified by traveler’s catchment area shows regionality in locations visited. (B) Campsite locations, colored by the number of households logged at a given location to show areas commonly visited. Satellite image from Leaflet package in R, sourced by Esri. See **S1 Fig** for maps with all points (day and campsites) logged.

At the individual traveler level, four travel patterns emerged from the GPS data (**Fig 4**, **S2-5 Fig**). The most common trip type was Long Term with 41.4% (24/58) of travelers, followed by Transient (34.4%, 20/58), Static (19.0%, 11/58), and Day trips (5.2%, 3/58) (**Table 1**). As expected, travelers with Static trips had the fewest unique campsites logged (1, IQR 1-2) and covered the smallest distance over the duration of the trip (a median total distance of 33.5 kms, IQR 23.0-54.9). With a median duration of 70 days, the Static trip durations were generally longer than the other three trips (~55 days). At the other end of the spectrum, travelers with Transient trips logged the most unique campsites (17, IQR 11-32.8) and traveled the furthest (278.5 km, IQR 186.3-557.4). While travelers conducting Long Term trips logged more unique nighttime locations than Day trips (10.5 vs 1), they logged fewer kilometers on average than Day trips (106 vs 157km). Of the four female travelers carrying GPS loggers, three were recorded conducting Long Term trips and one a Static trip (**Table 1**).

**Fig 4.**
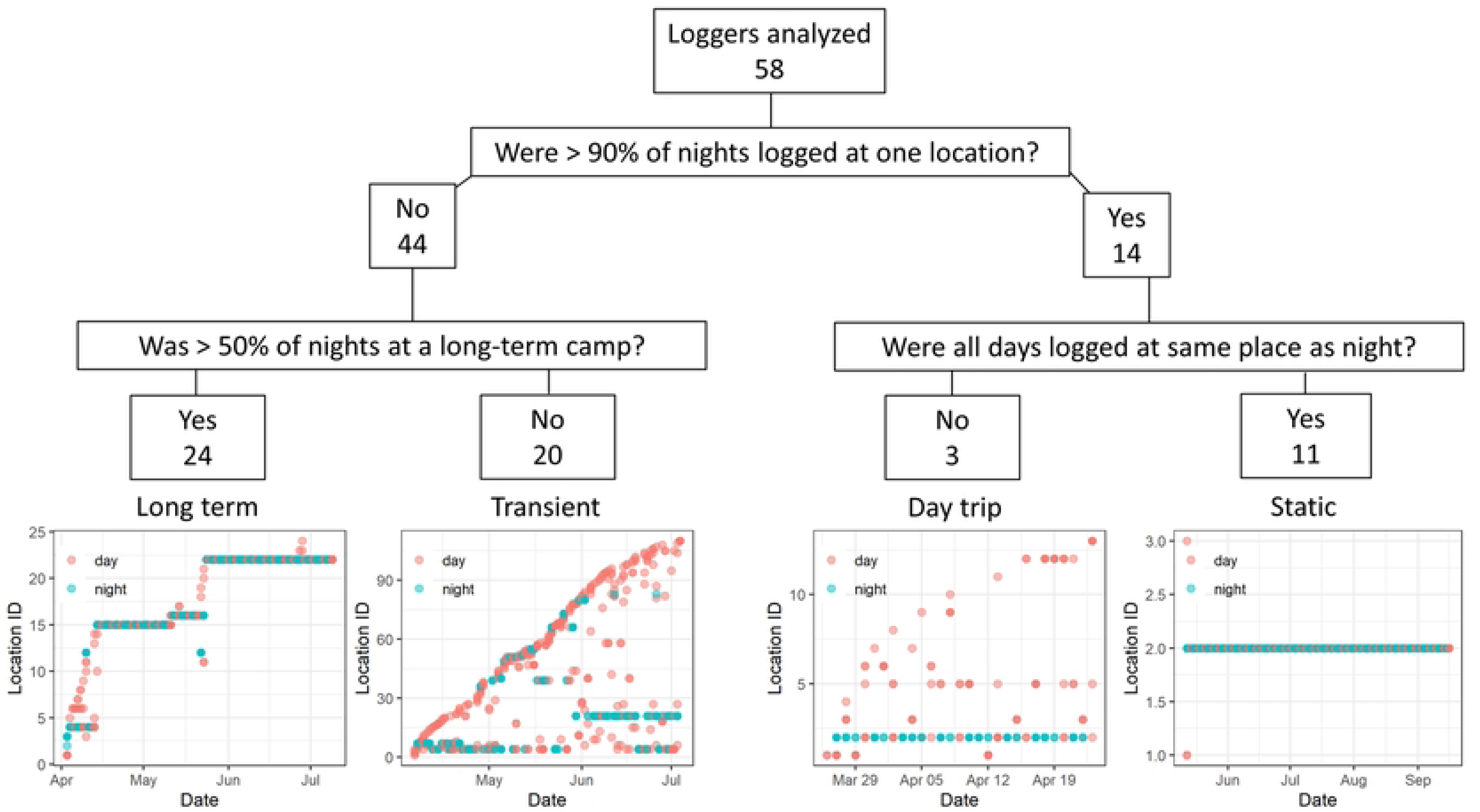
Individual trip patterns from GPS logger data. Using GPS logger data, we defined trajectories based on night (blue) and day (red) locations logged by each traveler. Both Long Term and Transient travelers logged a variety of night and day locations; however, Long Term travelers tended to spend ≥ 7 consecutive nights at each campsite whereas Transient travelers tended to spend < 7 consecutive nights at a campsite. Day trip and Static travelers both spent > 90% of their nights at the same location. They differed by the way Day trip travelers visited different locations during the day, while Static travelers logged all night and day points at the same location. The bottom four plots are tracks from four individuals that exemplify the different types of trip patterns. Tracks from all travelers are found in **S2-5 Fig**.

To further characterize these trip patterns, travel history from the surveys was incorporated. For most trip types, travel surveys tended to underestimate the number of campsites calculated from the GPS loggers (**Table 1**) and did not distinguish between the distances covered (i.e., using travel time to a camp as a proxy for distance). These differences possibly reflect recall bias and suggest that a travel survey alone might not detect the nuances of different trip types. The travel surveys collected from travelers who conducted Static trips indicated that these travelers visited 1 campsite which took 2-3 days to reach, suggesting that these travelers may have left their GPS loggers at their homestead while they traveled with the herd (**Table 1**); however, this cannot be confirmed. The median age was similar for Long Term, Transient, and Static trips (36-38 years), but was lower for Day trips (26 years). The majority of travelers for each trip type reported non-household members near their campsites, with a median of 4-6 people reported as near the campsites for all trip types. The majority of travelers on all trip types reported traveling with goats (≥95) and sheep (≥80%) and having an open water source near their camp (≥ 83.3%). Closed water sources were reported the most by Long Term travelers (42.7%) and the least by Transient travelers (20%).

### Malaria study population

To understand how traveling with the herd might impact infectious disease exposure, we compared the prevalence of malaria in travelers with that of remainers. From the 250 participating households, a total of 929 members were enrolled, consisting of the 304 members who planned to travel and 625 members who planned to remain. At follow-up, 18 travelers and 93 remainers were lost to follow-up, 70 people who planned to travel ended up remaining, 37 people who planned to remain ended up traveling, and 7 new travelers and 30 new remainers joined the study, resulting in 260 traveling members and 595 remaining members surveyed after seasonal travel **(Fig 2)**. Our goal was to measure infections acquired during the travel period so we only included those *without* malaria at baseline in the subsequent analysis. We excluded individuals with no baseline data (n=94), incomplete PCR results at either timepoint (enrollment n=15, follow-up n=7), and malaria at baseline by PCR (n=68). This resulted in 218 travelers and 453 remainers from 242 households in the analysis sample.

From the analyzed cohort, there was a median of 3 members (IQR: 3-5), 1 traveler (IQR 1:1) and 2 remainers (IQR 1:3) enrolled per household (**Table 2**). At least one child ≤15 years old was enrolled for 59.4% (142/239) households. The majority of households reported using open water sources for drinking and cooking (65.7%, 157/239), and relied on livestock as their primary source of income (63.2%, 151/239). All households reported ownership of livestock, with most owning goats (99.6%, 237/238) and sheep (93.7%, 223/238). All households reported they had no available bednets.

**Table 2.** Household characteristics of members included in the malaria analysis. % (number/N); median (IQR)

|  | Household<br>(N = 239) |
| --- | --- |
| People per household enrolled | 3 (3-5) |
| Travelers per household enrolled | 1 (1-1) |
| Remainers per household enrolled | 2 (1-3) |
| At least 1 child ≤ 15 years enrolled | 59.4 (142/239) |
| Number of nets per household | 0 (0-0) |
| **Catchment area** |  |
| Kangirisae | 25.5 (61/239) |
| Kerio | 25.5 (61/239) |
| Louwae | 23 (55/239) |
| Nakurio | 25.9 (62/239) |
| **Nearby Water source\*** |  |
| Only open | 65.7 (157/239) |
| Only closed | 45.2 (108/239) |
| **Primary income source** |  |
| Livestock | 63.2 (151/239) |
| Burning Charcoal | 33.1 (79/239) |
| Weaving | 17.2 (41/239) |
| Farming | 1.7 (4/239) |
| Informally Employed<sup>1</sup> | 2.1 (5/239) |
| **Households own certain livestock** |  |
| Goats | 99.6 (237/238) |
| Sheep | 93.7 (223/238) |
| Camels | 26.8 (64/239) |
| Cattle | 8.0 (19/239) |

<sup>1</sup>Informally Employed: Income from a relative or working at a small business; \*See Table 1 footnotes for water source details

Remainers tended to be female (67.3%, 305/453) with a median age of 19 years (IQR: 10-32), relative to travelers who were predominantly male (86.2%, 188/218) with a median age of 30.5 years (IQR: 21.3-42.0) (**Table 3**). Remainers included a larger proportion of children 15 years and younger (Remainers: 41.3%, 187/453, Travelers: 14.2%, 31/218), while travelers had a larger proportion of adults > 40 years (Remainers: 11.5%, 52/453, Travelers: 26.6%, 58/218). Most remainers (96.5%, 437/453) and travelers (93.1%, 203/218) did not report any symptoms at follow-up and few reported being recently sick, although travelers were twice as likely to report being sick since enrollment (Travelers: 3.7%, 8/218, Remainers: 1.8%, 8/452). Of those who reported being recently sick, 75% (6/8) of travelers self-medicated (i.e., took medicine or herbs from home or bought medicine from a pharmacy) while 75% (6/8) of remainers tended to visit a health facility. Of the participants who tested for malaria when feeling sick, travelers had a higher reported malaria positivity rate (Travelers: 100%, 3/3, Remainers: 50%, 1/2), although the number of observations was very small. Travelers who reported feeling recently sick reported taking medicine less often than remainers (Travelers: 50%, 4/8, Remainers: 75%, 6/8).

**Table 3.** Characteristics of travelers and remainers. (all and those who acquired malaria during the migration window) % (number/N); median (IQR)

|  | Traveler |  | Remainer |  |
| --- | --- | --- | --- | --- |
|  | All<br>(N = 218) | PCR+<br>(N = # in category) | All<br>(N = 453) | PCR+<br>(N = # in category) |
| **Gender** |  |  |  |  |
| Female | 13.8 (30/218) | 10.0 (3/30) | 67.3 (305/453) | 3.3 (10/305) |
| Male | 86.2 (188/218) | 9.0 (17/188) | 32.7 (148/453) | 6.8 (10/148) |
| **Age (yrs)** |  |  |  |  |
| ≤15 | 14.2 (31/218) | 6.5 (2/31) | 41.3 (187/453) | 6.4 (12/187) |
| 16-40 | 59.2 (129/218) | 8.5 (11/129) | 47.2 (214/453) | 3.3 (7/214) |
| >40 | 26.6 (58/218) | 12.1 (7/58) | 11.5 (52/453) | 1.9 (1/52) |
| Median age | 30.5 (21.3-42.0) | 31.5 (20.5 - 46.0) | 19.0 (10-32) | 12.0 (6 -26) |
| **Catchment area** |  |  |  |  |
| Kangirisae | 23.9 (52/218) | 9.6 (5/52) | 19.4 (88/453) | 4.5 (4/88) |
| Kerio | 25.7 (56/218) | 8.9 (5/56) | 26.5 (120/453) | 5.8 (7/120) |
| Louwae | 25.7 (56/218) | 7.1 (4/56) | 28.0 (127/453) | 3.1 (4/127) |
| Nakurio | 24.8 (54/218) | 11.1 (6/54) | 26.0 (118/453) | 4.2 (5/118) |
| **Water source since enrollment\*** |  |  |  |  |
| Open | 87.2 (190/218) | 8.4 (16/190) | 65.1 (295/453) | 5.1 (15/295) |
| Closed | 30.7 (67/218) | 11.9 (8/67) | 48.6 (220/453) | 3.2 (7/220) |
| **Medical History** |  |  |  |  |
| No symptoms on day screened | 93.1 (203/218) | 7.9 (16/203) | 96.5 (437/453) | 4.1 (18/437) |
| Sick since enrollment | 3.7 (8/218) | 50.0 (4/8) | 1.8 (8/452) | 12.5 (1/8) |
| Action taken when sick |  |  |  |  |
| Visited health facility | 25.0 (2/8) | 50.0 (1/2) | 75.0 (6/8) | 16.7 (1/6) |
| Self-medicated | 75.0 (6/8) | 50.0 (3/6) | 12.5 (1/8) | 0.0 (0/1) |
| Took malaria test | 37.5 (3/8) | 100.0 (3/3) | 25.0 (2/8) | 50.0 (1/2) |
| Positive malaria test | 100.0 (3/3) | 100.0 (3/3) | 50.0 (1/2) | 100.0 (1/1) |
| Took medicine | 50.0 (4/8) | 75.0 (3/4) | 75.0 (6/8) | 16.7 (1/6) |
| Antimalarial | 37.5 (3/8) | 100.0 (3/3) | 12.5 (1/8) | 100.0 (1/1) |
| Antibiotic | 12.5 (1/8) | 0.0 (0/1) | 62.5 (5/8) | 0.0 (0/5) |
| Pain Killer | 50.0 (4/8) | 75.0 (3/4) | 75.0 (6/8) | 16.7 (1/6) |

279 \*See Table 1 footnotes for water source details

### Malaria analysis

After travelers returned, 9.2% (20/218) of travelers (9.3%, n=16/172 of non-GPS carriers and 8.7%, n=4/46 of GPS carriers) and 4.4% (20/453) of remainers tested positive for malaria by PCR (**Table 3**). Acquisition of new infections was higher in travelers than remainers across gender, age group, catchment area, and type of water source (open or closed). While infection was similar for both male and female travelers (9-10%), it was twice as high in male remainers (6.8%, 10/148) than female remainers (3.3%, 10/305). Children ≤ 15 years had similar malaria burdens, regardless of their travel status (Travelers: 6.5%, 2/31; Remainers: 6.4% 12/187); however, new malaria infections increased with age for travelers (up to 12.1% in >40-year-olds) and decreased with age for remainers (down to 1.9% in >40-year-olds). The catchment area with the lowest number of infections was Louwae for both groups (7.1% (4/56) of travelers and 3.1% (4/127) of remainers) while the catchment areas with the highest was Nakurio for travelers (11.1% (6/54)) and Kerio for remainers (5.8% (7/120)). There was one month where malaria was more common in remainers followed up in that month than in returning travelers; otherwise, malaria infection in travelers was similar to or greater than the prevalence in remainers in all the study months (**S6 Fig**). Newly acquired infections reached a maximum of 15.5% (13/84) in travelers returning in July and 8.3% (2/24) in remainers after trips concluding in May.

Individual-level models identified characteristics associated with acquiring an infection over the course of traveling with the herd (**Table 4**). Univariate analysis revealed that the odds of getting malaria was more than two times greater in travelers (aOR=2.19, 95% CI: 1.15-4.18) relative to remainers and in males (aOR = 2.16, 95% CI: 1.11 - 4.40) relative to females. Age group did not appear to be a statistically significant factor. When adjusting for travel status, age and gender, travel status and gender were no longer statistically significant; however, there were a relatively small number of infections which limited the power of the study. The general trend suggests gender and travel are still associated with an increased odds of malaria (aOR=1.91 (0.86-4.32) for travelers relative to remainers and aOR = 1.54 (0.70-3.48) for males relative to females). While trip types could not be characterized for all travelers and thus was too sparse a factor to include in the model, it is interesting to note that new malaria infection in GPS carriers was 0% after Static (0/10) or Day trips (0/2), 5.6% (1/18) after a Transient trip, and 13.6% (3/22) after a Long Term trip.

**Table 4.**
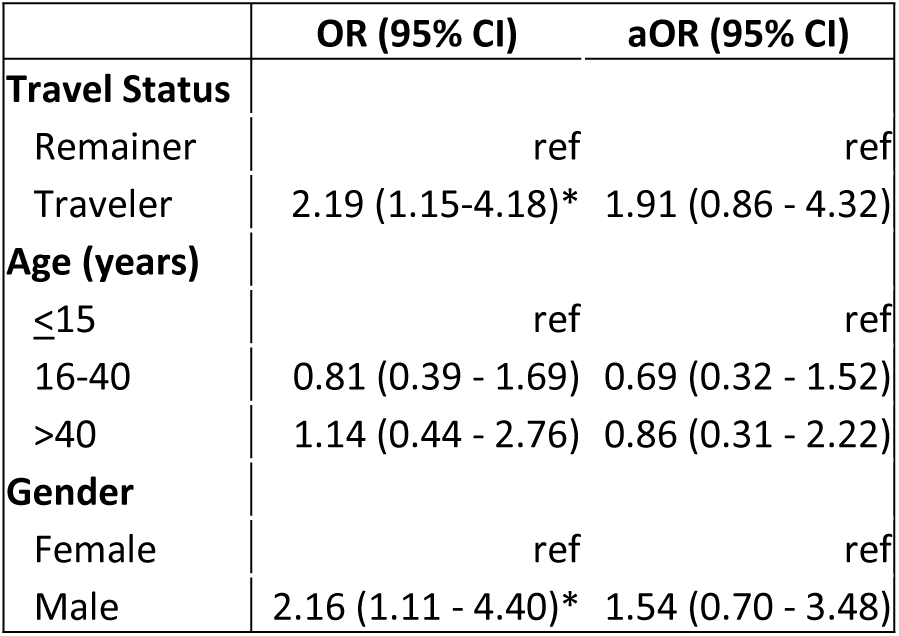
Factors associated with PCR+ malaria cases. Significance levels at or below 0.05 are indicated by a*.

|  | OR (95% CI) | aOR (95% CI) |
| --- | --- | --- |
| **Travel Status** |  |  |
| Remainer | ref | ref |
| Traveler | 2.19 (1.15-4.18)* | 1.91 (0.86 - 4.32) |
| **Age (years)** |  |  |
| ≤15 | ref | ref |
| 16-40 | 0.81 (0.39 - 1.69) | 0.69 (0.32 - 1.52) |
| >40 | 1.14 (0.44 - 2.76) | 0.86 (0.31 - 2.22) |
| **Gender** |  |  |
| Female | ref | ref |
| Male | 2.16 (1.11 - 4.40)* | 1.54 (0.70 - 3.48) |

## Discussion

Understanding the relationship between mobile communities’ travel patterns, health-care seeking behaviors, and risk for disease transmission is critical for informing intervention strategies suitable for their lifestyles and aiding in broader disease elimination campaigns. Here, we quantified travel patterns in four semi-nomadic communities and determined that the acquisition of new malaria infections was twice as high in individuals who traveled with their herd than household members who remained behind. While malaria was acquired at similar proportions for male and female travelers, it was twice as high in males than female remainers, suggesting that gender norms may play a role in exposures around the homestead. Travel patterns in these areas had not been well characterized and we describe local movement of herders that is quite heterogeneous within a small geographic area. Although direction of travel is more similar amongst individuals within a catchment than between, the distance, duration and overnight destinations were highly variable across individuals. None of the households reported access to a bednet and few travelers sought treatment from a health facility when they felt sick, consistent with the tendencies for mobile communities in general to have reduced access to prevention and health care(12,39). Community-based strategies, where individuals are taught how to detect and manage infections in their communities when access to healthcare is limited, have been successful in other nomadic communities(12); however, many of this study’s participants who tested positive for malaria were asymptomatic which would make it challenging to know when to take action. Additionally, mobile clinics placed along well traveled routes have been used to provide health care to patients who would not have had access otherwise(40); however, this study did not reveal any common routes in these communities which would make it difficult to determine where to place a mobile clinic. Screening travelers upon their return would be a proactive approach, but likely unrealistic, given how often travel occurs, the unpredictable nature of when and where the trips will take place, and the resource constraints on intensifying control efforts. Instead, focusing on preventative measures, such as bednet distribution in more remote areas would be conducive for reducing malaria transmission in travelers who spend most of their nights at the same place (i.e., Day trip or Static trips) and also prevent onward transmission from infected travelers upon their return. For travelers who spend more nights away from their settlements (i.e., Long term or Transient travelers), there is a need for solutions that would easily integrate into their lifestyle, such as insecticide treated clothing(41). Ultimately, maintaining an open dialogue with mobile communities about their needs and co-developing practical solutions is critical to ensure interventions are maximally useful.

While this study has established that travel patterns can be very heterogeneous both in duration and destination within communities and that malaria is often acquired while traveling with the herd, a number of important research areas remain. First, there is a need to define the transmission dynamics within and between settled and mobile communities. From previous work, we know malaria is endemic in the neighboring settled communities in central Turkana(35); however, the role mobile communities play in local and regional circulation needs to be defined. Second, we still do not fully understand where the travelers were acquiring malaria on their travels. GPS data did not reveal potential transmission hotspots and the satellite imagery accessed in the analysis did not have the spatial nor temporal resolution to pick up on transient water sources travelers may have stayed near. A third aspect that needs to be elucidated is the interaction between pastoralists, their herds and their risk of malaria. If the mosquitoes in the area are opportunistic feeders, they may be attracted to feed on large herds and bite humans nearby(42). Finally, this study only focused on a few semi-nomadic communities in Central Turkana. Movement in Central Turkana is less extensive than among nomadic groups in Western Turkana, where households often travel hundreds of kilometers with their entire family and cross into Uganda or Ethiopia (personal communication with local health authorities). Additional studies are needed to characterize these movements, exposures, and health seeking behaviors before conclusions can be drawn. However, based on our findings, we suspect that those who travel with their herds will still have higher risks of malaria exposure. More investigation is needed to better understand transmission dynamics and opportunities for intervention in this unique context.

The limitations of this study include challenges with collecting complete information at enrollment and follow-up, which ultimately lead to a smaller sample size. In addition, it was difficult to know whether the GPS loggers were actually being carried. For instance, the fact that 11 loggers recorded points for multiple weeks within the same 0.5km^2^ area suggests that they might have been left behind. While this may have led to mis-categorization of some trips, we were still able to categorize three different trip patterns that would be informative for different intervention approaches. Most of the variables we explored were self-reported, therefore we cannot rule out mis-reporting things like nets, livestock, and symptoms, or recall bias for trip details and recent illnesses. Finally, we only screened for *P. falciparum;* however, recent literature/preliminary data suggests that *P. vivax* is also circulating in this region(43). If this is true, then the prevalence of malaria in this study (and the region in general) is being under-reported.

In conclusion, this study is one of the few that quantifies mobility patterns of and defines the relationship between mobility patterns and disease exposure in mobile communities. We determined that traveling with the herd doubled the odds of acquiring new malaria, relative to those who remained behind. The four different travel patterns identified could be used to inform intervention strategies more suitable to a mobile life-style. Further studies are needed to determine how these observations can be generalized to other disease exposures as well as the role of mobile populations on disease transmission with the broader community.

## Data Availability

The data will be deposited in Duke University Digital Research Data Repository prior to publication

## Acknowledgements

We thank our field team, especially Dennis Okoth, Rose Adome, Erastus Lomuria, Benson Lorunye, Ekapeton Jackline, Lorinyo Francis Ethuron, Esinyen Mark, Topuye Ekuom David, Akolonyo Peter Ikao, James Lomaala, Anjeline Atabo, Bosco Nawoto, and all the families who gave their valuable time to participate in this study.

## Supporting Information

**S1 Text and Table. Using Reported departure and return dates instead of the enrollment and follow-up dates**

**S2 Text and Table. Sensitivity analysis around definition of “long term campsite”**

**S1 Fig. Population level trip characteristics**. (A) All locations logged, stratified by traveler’s catchment area shows regionality. (B) Campsite locations, colored by the number of households logged at a given location to show areas commonly visited. Satellite image from Leaflet package in R, sourced by Esri.

**S2 Fig. GPS carrying travelers whose trips were categorized as Static.** >90% of the night spots were spent at the same night location they were enrolled at, but most day points were logged at different locations.

**S3 Fig. GPS carrying travelers whose trips were categorized as Day trips**. >90% of the night spots were spent at the same location and most night and day points were logged at the same location.

**S4 Fig. GPS carrying travelers whose trips were categorized as Transient.** <90% of the night spots were spent at the same location and > 50% of nights were spent at transient camps (defined as camps with < 7 consecutive nights spent).

**S5 Fig. GPS carrying travelers whose trips were categorized as Long Term.** <90% of the night spots were spent at the same location and > 50% of nights were spent at long term camps (defined as camps with >7 consecutive nights spent).

**S6 Fig. Proportion of remainers and travelers who tested PCR+ for malaria after the migration period, stratified by the month the travelers returned.**

